# School teachers’ self-reported fear and risk perception during the COVID-19 pandemic – a nationwide survey in Germany

**DOI:** 10.1101/2021.06.17.21258956

**Authors:** Stefanie Weinert, Anja Thronicke, Maximilian Hinse, Friedemann Schad, Harald Matthes

**Affiliations:** Institute of Social Medicine, Epidemiology and Health Economics, Charité-Universitätsmedizin Berlin, corporate member of Freie Universität Berlin, Humboldt-Universität zu Berlin, and Berlin Institute of Health, Berlin, Germany; Research Institute Havelhöhe, Kladower Damm 221, 14089 Berlin, Germany; Interdisciplinary Oncology and Palliative Care, Hospital Gemeinschaftskrankenhaus Havelhöhe, Berlin, Germany; Medical Clinic for Gastroenterology, Infectiology and Rheumatology CBF, Institute of Social Medicine, Epidemiology and Health Economics, Charité-Universitätsmedizin Berlin, corporate member of Freie Universität Berlin, Humboldt-Universität zu Berlin, and Berlin Institute of Health, Berlin, Germany

**Keywords:** COVID-19, vaccination intention, fear of infection, teacher, schools, survey

## Abstract

**Background:** With COVID-19 cases peaking, COVID-19 vaccination programs starting and health systems reaching their limits in winter 2020/21, schools remained closed in many countries despite ever-recurring debates. To better understand teachers’ fear of infection and risk perception we conducted a survey in Germany.

**Methods:** Participants were recruited through various associations and invited to take part in a cross-sectional COVID-19 specific online survey. Anonymous demographic and self-reported data were collected from those who gave their informed consent. Descriptive statistical analysis was performed. To evaluate with fear associated factors of contracting SARS-CoV-2, an adjusted multivariable regression analysis was performed.

**Results:** 6.753 teachers gave their informed consent to answer the online survey. The median age of the teachers was 43 years with 77% being female. Most teachers worked in high schools (29%) and elementary schools (26%). Most participants (73%) feared to contract SARS-CoV-2 at school while 77% intended to be vaccinated against COVID-19. 98% considered students to pose the greatest risk. Multivariable regression analysis revealed that female and younger teachers were significantly more anxious to get infected with SARS-CoV-2 and that the odds teachers were more anxious was 9 times higher for those who favored re-opening of schools the least (p < 0.001).

**Conclusions:** To the authors’ knowledge, this is the first study to describe teachers fear and risk perception of COVID-19 and their attitude towards vaccinations in a nationwide survey. The anxiety correlates to the COVID-19 protection measures demanded. Teachers’ fear is the driving factor and not a rational logic.

## Introduction

In the pandemic era of COVID-19, the scientific interdisciplinary discourse on various coping strategies has largely been conducted by policy makers and their advisors. This has resulted in many opinions and views that often differ from the scientific facts. In fact, more than 3 million people in Germany have been tested positively for SARS-CoV-2 so far. Nearly 80,000 deaths are linked to Corona Virus Disease 2019 (COVID-19) (as of April 14, 2021, (1)).

Prior to the launch of the vaccination campaign in November 2020, the German population’s intention to get vaccinated was 37% (2). In the events of the second and third waves and with increasingly stricter COVID-19 measures in place, the vaccination intention rose to 59% in February and 75% in May 2021. However, these are overall numbers that hardly mirror the vaccination intention of different occupational groups which may have an above average risk of SARS-CoV-2 infections. Intensive care healthcare workers’ vaccination intention had risen from 65% to more than 75% between December 2020 and February 2021 (3). A survey conducted in Saxony, Germany at about the same time showed that a staggering 90% of teachers from public schools and around 80% in private school settings intended to receive COVID-19 vaccinations (4). A reason for this and whether this applies nationwide is unknown. Healthcare workers are clearly at particularly high risk of contracting SARS-CoV-2 through physical contact to infected people. Teachers are also at high risk. However, as classroom teaching has been largely reduced and COVID-19 precautions have been implemented in schools, most teachers are to a certain extent protected. The actual risk of teachers contracting SARS-CoV-2 despite COVID-19 precautions remains controversial.

Children and adolescents probably spread the infection at similar rates (5). While at the beginning of the pandemic studies observed no evidence of secondary transmission of COVID-19 from children attending school (6), it is now clear that infections have been imported into schools from the community where it can spread (7). But further transmission within schools has been rare when rigorous measures such as wearing face masks and frequent ventilation of rooms have been implemented (8–10) suggesting that schools do not substantially contribute to increased circulation of SARS-CoV-2 among local communities.

However, the actual risk and the risk perception of teachers may differ significantly. With the multiwave pandemic dynamics in winter and spring 2020/21 hitting most countries hard, with health care systems reaching theirs limits and with schools remaining to be closed, we wanted to better understand teachers’ fear of infection and risk perception. Therefore, we invited teachers in Germany to participate in the newly developed ImpfREAD survey, in which we asked teachers’ fears and concerns contracting SARS-CoV-2, their intention to be vaccinated against COVID-19, and their opinions on new virus variants and COVID-19 fighting strategies discussed by policy makers. Demographic and medical parameters were also collected to assess the risk of suffering severe COVID-19. The survey was conducted once and online.

## Methods

### Study design

We conducted a cross-sectional real-world anonymous survey to determine teachers’ fear contracting SARS-CoV-2 and their risk perception, their intention to receive COVID-19 vaccinations, and their opinion on COVID-19 precautions. The survey was conducted nationwide in Germany. Data collection took place between March 2 and April 30, 2021.

### Participants and enrolment

Participants were recruited through the Education and Training Association (VBE, Verband Bildung und Erziehung), German Teachers’ Association (DL, Deutscher Lehrerverband), Union for Education and Training (GEW, Gewerkschaft für Erziehung und Bildung), Association of German Private School Associations (VDP, Verband Deutscher Privatschulverbände e.V.), Association of Waldorf Schools (Bund der Freien Waldorfschulen) and the Montessori Association (Montessori Dachverband). The link to the survey was distributed to association members via email distribution lists. The educational network News4Teacher featured the survey on its website. School boards which were contacted directly also distributed the link via distribution lists. The Twitter account of the Institute for Social Medicine, Epidemiology and Health Economics (@CSozialmedizin) posted the survey in a newsfeed. A snowballing system was encouraged, asking participants to share the survey with other teachers.

The study was conducted in accordance with the ethical standards of the 1964 Declaration of Helsinki. Since the data in our study was collected anonymously (no code lists available, no conclusion could be drawn from the collected data to personal data) ethical approval was waived. The data collection took place in accordance with the European Data Protection Regulation. Prior to participating in the survey, participants were given a description and objectives of the study. Written informed consent has been obtained from all the participants prior study enrolment. The study was approved by the institutional data protection board and quality management based on the given data protection concept.

### Survey and Outcomes

The survey was newly developed in a multi-disciplinary research collaboration between life scientists, psychologists, and physicians. The primary outcome was to clarify the impact of the COVID-19 outbreak on teacher’s individual’s health-related worries and their intention to receive COVID-19 vaccinations. The survey consisted of 9 parts capturing a) socio-demographic characteristics, such as age, gender and school type, as well as teachers’ fear of contracting SARS-CoV-2, b) teachers’ intentions to get vaccinated, c) prioritization of vaccines distribution and school re-openings, d) teachers’ fear of suffering severe COVID-19, e) teacher’s opinion on compulsory and f) influenza vaccinations, g) teachers’ concern about new variants and the third COVID-19 wave, h) teachers conspiracy mentality, and i) socio-demographic characteristics such as numbers of children and underlying diseases. The ImpfREAD survey consisted of a total of 92 items. Depending on the item participants could either indicate how likely they agree to each item on a five-point Likert scale ranging from ‘I agree absolutely’ to ‘I don’t agree at all’, answer “yes” or “no” or choose from given answers.

### Data collection

Data collection was performed by means of a structured, anonymous, self-administered questionnaire (Appendix 1) using the Research Electronic Data Capture (REDCap) data management platform version 10.6.14 hosted at Charité-Universitätsmedizin Berlin. REDCap is a secure web application for building and managing online surveys and databases (11). Patients with missing data were not included.

### Statistical analyses

Continuous variables were described as median with interquartile range (IQR); categorical variables were summarized as percentages. Univariate data was analyzed using MS Excel 2000 and GraphPad Prism version 9.0.0. To address potential sources of bias, an adjusted multivariable regression analysis with dichotomic outcome of anxiousness of getting COVID-19 (yes/no) was performed using the software R (Version 3.6.1, R Development Core Team, Vienna, Austria). p-values < 0.05 were considered to be significant.

## Results

### Participants

The online survey was accessed by 6.995 individuals. 242 individuals either did not give informed consent or provided any information. 6.753 teachers providing their informed consent were included in the study and records were analyzed. Most of the participants (42%) were recruited through the educational magazine News4Teacher, via VBE and GEW (33%), school boards (17%), and other sources (8%) (Supplementary table S1).

### Demographic data

The study was conducted with 6.753 teachers in Germany. The geographical distribution of the participating teachers was rather homogeneous with most participants coming from northern Germany (Supplementary table S2). Table 1 lists the socio-demographic characteristics of the participants. The median age of the teachers was 43 years (IQR: 36-51 years) and 77% were female. Most participants worked in high schools (29%) and elementary schools (26%). 7% taught in private school settings (Waldorf schools, Montessori schools, private schools). Most of the participating teachers (40%) taught grades 7 – 13 students with German language (49%) and mathematics (42%) as the main subjects. Many of the teachers (64%) had children, with having two children (48%) or three or more children (22%). 13% were single parents. Almost 5% of the teachers reported to have had COVID-19 and 16% had been vaccinated against COVID-19. 29% reported to have one or more pre-existing conditions, e.g., 18% reported to have asthma or any other respiratory disease or hypertension and 2% being obese.

**Table 1.** Socio-demographic data of the participating teachers.

| Variable | n (%) |
| --- | --- |
| **Gender (n=5894)** |  |
| Female | 4513 (76.6) |
| Male | 1358 (23.0) |
| Divers | 23 (0.4) |
| **School types (n=5876)** |  |
| Elementary schools | 1510 (25.7) |
| Secondary schools (Hauptschulen)<sup>#</sup> | 212 (3.6) |
| Secondary schools (Realschulen)<sup>#</sup> | 708 (12.0) |
| High schools (Gymnasien)<sup>#</sup> | 1729 (29.4) |
| Comprehensive schools (Gesamtschulen)<sup>#</sup> | 943 (16.0) |
| Schools for children with special needs | 344 (5.9) |
| Waldorf schools | 223 (3.8) |
| Montessori schools | 23 (0.4) |
| Boarding schools | 7 (0.1) |
| Private schools | 151 (2.6) |
| Language schools | 26 (0.4) |
| **Grade levels (n=5525)** |  |
| Grades 1 – 6 | 1658 (30.0) |
| Grades 7 – 13 | 2191 (39.7) |
| All grades | 1676 (30.3) |
| **School subject (n=5532)** |  |
| German language | 2735 (49.4) |
| Mathematics | 2316 (41.9) |
| Science | 1490 (26.9) |
| Science (at elementary schools) | 1556 (28.1) |
| Foreign languages | 1708 (30.9) |
| Sports | 1034 (18.7) |
| Music | 874 (15.8) |
| Others | 3235 (58.5) |
| **Age (n=5904)** | Median: 43 years<br>IQR: 36-51 years |
| **Having children (n=5583)** | 3596 (64.4) |
| **Number of children (n=2930)** | 1 child: 1052 (29.4)<br>2 children: 1731 (48.3)<br>3 children and more: 799 (22.3) |
| **Single parents (n=3497)** | 462 (13.2) |
| **From COVID-19 recovered (n=5779)** | 266 (4.6) |
| **COVID-19 vaccine received (n=5878)** | 949 (16.1) |
| **One or more pre-existing conditions (n=6753)** | 1929 (29.3) |
| Asthma and other respiratory diseases | 606 (9.0) |
| Hypertension | 584 (8.6) |
| Obesity | 160 (2.4) |
| Type 1 or 2 diabetes | 104 (1.5) |
| Hashimoto | 96 (1.4) |
| Cancer | 71 (1.1) |

<sup>#</sup>In Germany, Hauptschulen, Realschulen, Gesamtschulen and Gymnasien refer to secondary school forms of intermediate education, i.e., level 2 according to UNESCO's ISCED classification. Students enter these schools after receiving a teachers' recommendation or, alternatively, after passing an entrance examination.

### Teachers’ fears of contracting SARS-CoV-2 at school

A striking majority of the participating teachers (73%) feared contracting SARS-CoV-2 at work (Figure 1, Supplementary table S3). Only 11% of participants were not at all afraid of getting infected at work. Students (98%), younger (44%) and older colleagues (30%) were named as major thread of infection (Figure 1, Supplementary table S4). Interestingly, 77% reported that teachers also pose a risk to students (Supplementary table S4). A majority of the teachers (71%) feared that students might carry and transmit SARS-CoV-2 even though being asymptomatic (Figure 1, Supplementary table S3). Additionally, many teachers agreed that schools and students substantially contribute to spread SARS-CoV-2 in communities and schools, respectively (66% and 65%, respectively, Figure 1, Supplementary table S3).

**Figure 1.**
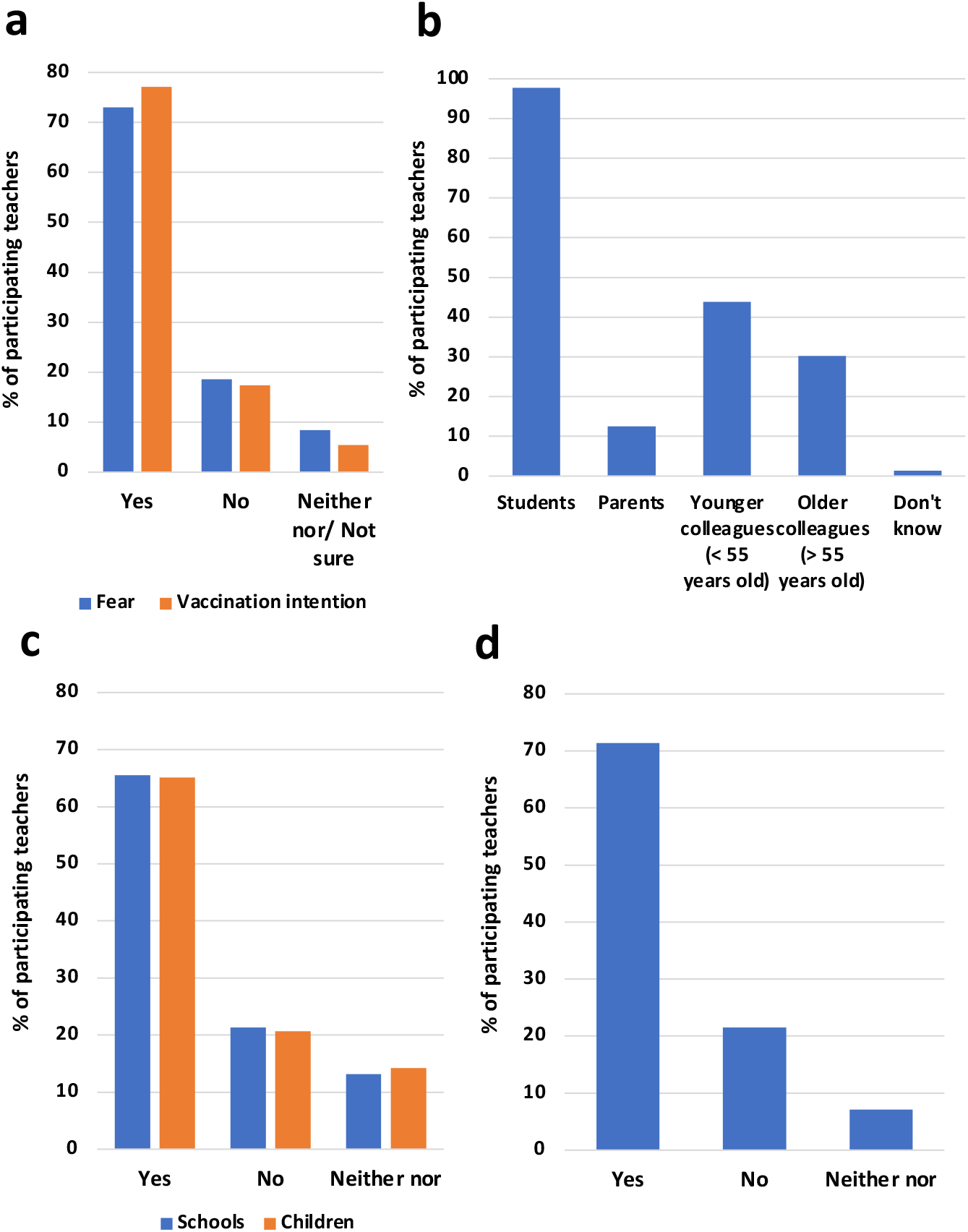
Teachers’ fear contracting SARS-CoV-2 (*n=5936*) and their intention to get vaccinated against COVID-19 (*n=5850*) are similarly high **(a)**. Teachers’ fear the most getting infected from students and younger colleagues (*n=4320*) **(b)** and believe that schools and children substantially contribute to circulation of SARS-CoV-2 (*n=5936*) **(c). (d)** Teachers’ fear that children can carry SARS-CoV-2 while being asymptomatic (*n=5930*).

Multivariable regression analysis adjusting for age, gender, intention to get vaccinated, school types, the opinion whether to open schools as highest priority, the risk perception of getting COVID-19, and the class subjects taught, revealed that female teachers (OR 1.92, p < 0.001) were significantly more anxious of getting COVID-19 compared to their male colleagues (Figure 2, Supplementary table S5). In contrast, teachers who don’t want or are unsure about getting vaccinated feared an infection significantly less (OR 0.02, p < 0.001, OR 0.17, p < 0.001, respectively) compared to teachers intending to get vaccinated. Interestingly, also teachers from Waldorf schools and schools for children with special needs showed significantly less fear (OR 0.38, p < 0.001; OR 0.63, p = 0.0264, respectively). Also, teachers who perceived their risk of getting COVID-19 as moderate, low, or not existing had significantly less fear (OR 0.12, p =0.000; OR 0.03, p < 0.001; OR 0.01, p < 0.001, respectively) compared to teachers who perceived their risk as high (Figure 2, Supplementary table S5).

**Figure 2.**
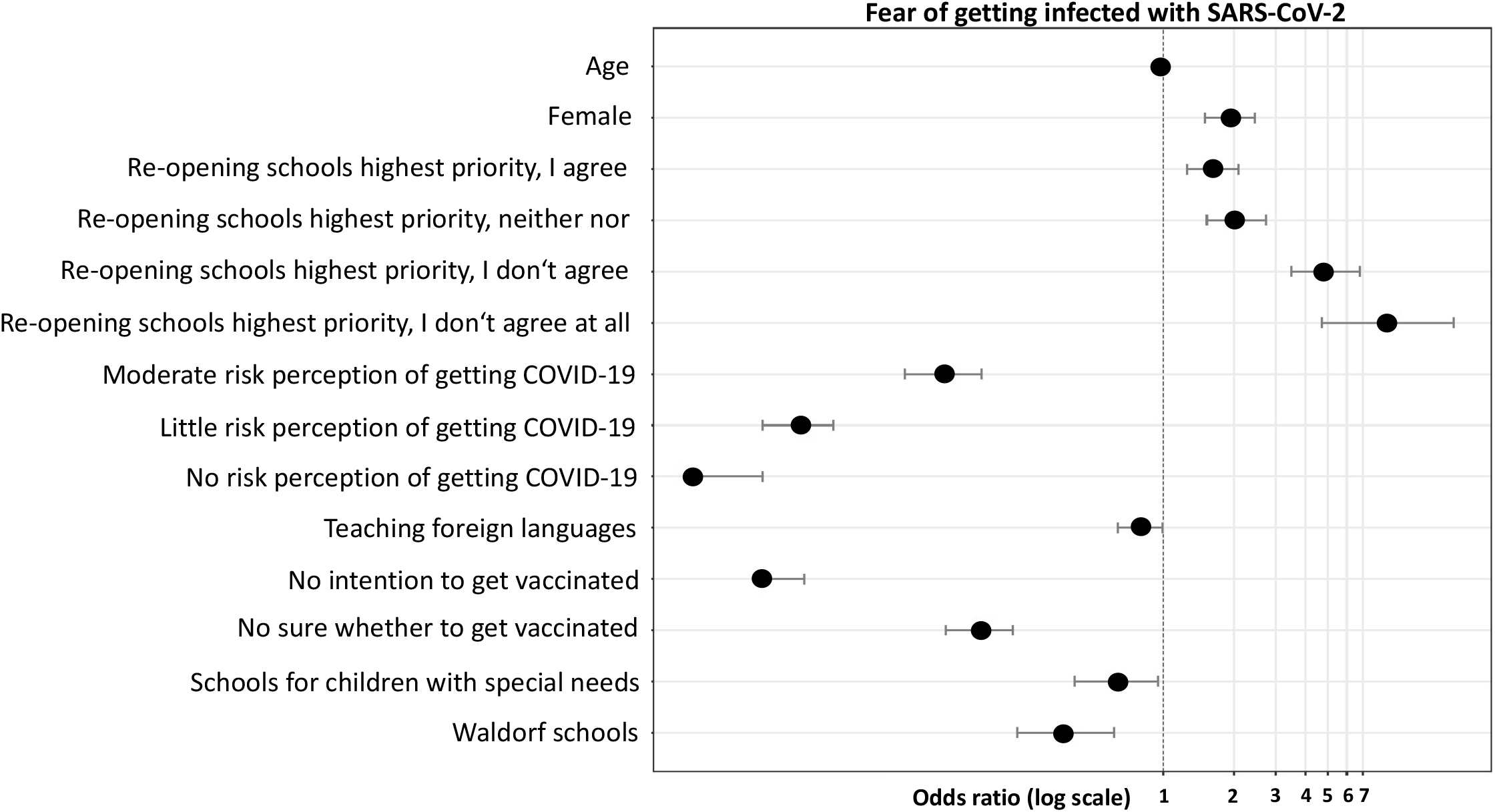
Probability of fear of getting infected with SARS-CoV-2, adjusted multivariable logistic regression analysis. Factors presented are significantly (p<0.05) associated with a reduced probability (left-hand side from the indicated margin) or with an increased probability (right-hand side from the indicated margin) to be anxious of getting infected with SARS-CoV-2.

Strikingly, teachers who oppose the re-opening of schools are 9 times more anxious than teachers who consider school re-openings as very important (OR 8.94, p < 0.001). At last, teachers of foreign languages were significantly less anxious than teachers not teaching foreign languages. With a tendency towards significance, music teachers were more anxious compared to non-music teachers (OR 1.34, p = 0.0501). Overall, the fear of COVID-19 decreases with age with the odds decreasing by 0.98 times per life year, i.e., younger teachers are significantly more anxious than older teachers (p = 0.0008).

Furthermore, we asked about possible conflicts with parents and teachers’ opinion on COVID-19 precautions. A majority (47%) felt that parents were too careless about SARS-CoV-2 and thought primarily of themselves and childcare (45%, Supplementary table S6). Most teachers (58%) reported to feel being under pressure to implement COVID-19 precaution state guidelines. Similarly, 49% felt they do not meet the expectations of parents and the states regarding the implementation of COVID-19 precautions while educating child-friendly and to a high standard (Supplementary table S6). Many teachers (45%) had given in at some point to parent’s insistence on additional COVID-19 precautions (Supplementary table S6).

### Teachers’ fear suffering severe COVID-19 and the new SARS-CoV-2 variants

35% of the participating teachers considered their risk of suffering severe COVID-19 as being increased, high or very high (Table 2). Many teachers volunteered to be tested for SARS-CoV-2 infections depending on the risk situation or at least once a week (42% and 33% respectively, Supplementary table S7).

**Table 2.** Teacher’s risk perception suffering severe COVID-19.

| Variable | n (%) |
| --- | --- |
| **"How large do you estimate your risk of suffering a server course of COVID-19?" (n=5784)** |  |
| Very little | 725 (12.5) |
| Little | 1336 (23.1) |
| Moderate | 1724 (29.8) |
| Elevated | 1230 (21.3) |
| High | 565 (9.8) |
| Very high | 204 (3.5) |
| **"I am concerned that due to the overload of the health care system, the options to go to a hospital and if necessary, an ICU are limited." (n=5801)** |  |
| I agree completely | 1470 (25.3) |
| I agree | 2276 (39.2) |
| Neither nor | 712 (12.3) |
| I don't agree | 735 (12.7) |
| I don't agree at all | 608 (10.5) |
| **"How do you judge the current non-pharmacological precautions (distance + hygiene + mask + warning app + ventilation)?" (n=5829)** |  |
| Exaggerated | 1003 (17.2) |
| Appropriate | 2565 (44.0) |
| Too little | 2139 (36.7) |
| I don't know | 122 (2.1) |
| **"My concerns about COVID-19 are getting stronger with the new corona mutations." (n=5726)** |  |
| I agree completely | 2111 (36.9) |
| I agree | 2035 (35.5) |
| Neither nor | 381 (6.7) |
| I don't agree | 531 (9.3) |
| I don't agree at all | 668 (11.7) |
| **"One should proceed with caution when re-opening schools, especially in view of the difficult-to-calculate dangers posed by new virus mutations." (n=5721)** |  |
| I agree completely | 2809 (49.1) |
| I agree | 1402 (24.5) |
| Neither nor | 377 (6.6) |
| I don't agree | 495 (8.7) |
| I don't agree at all | 638 (11.2) |
| **"I see a third corona wave approaching unnoticed due to the new variants." (n=5715)** |  |
| I agree completely | 2974 (52.0) |
| I agree | 1386 (24.3) |
| Neither nor | 407 (7.1) |
| I don't agree | 346 (6.1) |
| I don't agree at all | 602 (10.5) |

By the end of 2020, new variants of SARS-CoV-2 had spread from abroad to many European countries. These new variants increased concerns getting COVID-19 in 72% of the participating teachers (Table 2). A majority saw the third COVID-19 wave in Germany coming and was concerned that a strained health care system limits their ability to access a hospital and, if necessary, an intensive care unit (76% and 65%, respectively). Despite strict lockdown measures in place, 37% of the teachers rated the general non-pharmacological precautions as still too low (Table 2). A striking majority (78%) believed that COVID-19 can also become a seasonal illness due to mutations in SARS-CoV-2 (Supplementary table S8).

### Teachers’ opinion on vaccinations, vaccination strategy and priority setting

By the time of the survey period, only a minor fraction of the participating teachers (16%) had received a COVID-19 vaccination (Table 1). But most teachers (77%) intended to get vaccinated (Figure 1). Only 6% were unsure about the COVID-19 vaccination. Many teachers (72%) showed a strong preference for the BioNTech vaccine (Supplementary table S9). 43% and 22% of the participating teachers would also choose the Moderna and AstraZeneca vaccines, respectively. Surprisingly, 23% of the teachers did not have any preference towards a specific vaccine (Supplementary table S9). The main reasons given by most of the participants to get vaccinated were self-protection from getting COVID-19 (97%) and protection of family, friends, children, and others (92%, 75% and 78%, respectively) (Supplementary table S10). However, only 26% of the teachers choose returning to school quickly as a reason of getting vaccinated. On the other hand, teachers refrain from COVID-19 vaccinations for different reasons, such as unknown long-term effects (87%), vaccines not been adequately tested (78%), and concerns about adverse events (76%). Many participants felt a distrust of the manufacturers and vaccines (49% and 52%, respectively) (Supplementary table S10). Surprisingly, many teachers (67%) supported the notion that all colleagues should be vaccinated against COVID-19 before returning to work (Supplementary table S3).

In Germany, there are no compulsory vaccinations. However, in this study, teachers voted rather equally for and against a compulsory vaccination against COVID-19 discussed by policy makers (43% and 40%, respectively) (Supplementary table S11). Surprisingly, a majority (49%) supported the idea of compulsory vaccinations for other infectious diseases (Supplementary table S11).

A large majority (70%) has regularly checked that they had received all recommended vaccinations. In addition, almost 62% considered the recommendations of the Standing Committee on Vaccination (STIKO) to be appropriate (Supplementary table S12).

Until June 2021, COVID-19 vaccine allocation was subject to subdivision into priority groups by law in Germany. 71% of the participating teachers agreed with this division (Supplementary table S13). Similarly, 73% agreed that teachers receive the vaccination earlier when priority groups were changed in late February 2021. Furthermore, a majority (53%) rated re-opening schools as being a top priority (Supplementary table S13). However, regarding new virus variants and a third COVID-19 wave, more teachers (76%) wanted to proceed cautiously in re-opening schools (Table 2).

## Discussion

With the second COVID-19 wave hitting Germany at the end of 2020, schools became a focus of attention. Vaccinations had just begun but although teachers’ vaccination intention was above average and their role in society is highly acknowledged, they were not prioritized for vaccinations. A public debate arose around teachers’ vaccinations and the role of schools and children in spreading SARS-CoV-2. To investigate factors associated with teachers’ intention to be vaccinated against COVID-19 we conducted a cross-sectional survey of 6.753 teachers in Germany. Using the newly developed ImpfREAD survey, we aimed to investigate both fears and risk perceptions regarding a SARS-CoV-2 infection at school, as well as teachers’ attitudes towards vaccinations.

So far, only a few studies investigated teachers’ fear of contracting SARS-CoV-2. A cross-sectional study in summer 2020 in Berlin, Germany revealed that about half of the school staff showed medium to very strong fear of infection, and 59% reported moderate to very high perceived risk of infection (12). However, this study comprised only 112 teachers at 24 schools in Berlin, mirroring only a limited number of teacher’s opinion which in part can largely depend on their respective schools. Another study, conducted in February 2020 with nearly 9.000 teachers in three cities in China found a very low prevalence of anxiety of just under 14%, with women being slightly more anxious than men (13). Another study of almost 1.700 teachers conducted in September 2020 in Spain, reported that nearly half of the teachers surveyed had anxiety (14). However, both latter studies used anxiety scales which do not specifically focus on the situation during a pandemic. In our study we found 73% teachers being afraid of getting infected with SARS-CoV-2 at school. Female teachers were significantly more anxious than their male colleagues which is consistent with the fact, that women are twice as likely to be diagnosed with an anxiety disorder than men (15).

It is striking that teachers’ anxiety correlates with their attitude towards school re-openings, i.e., the less teachers rated school re-openings as a priority, the greater their anxiety was. Interestingly, teachers at Waldorf schools were significantly less anxious than teachers at primary schools. Rudolf Steiner’s concept of health strongly incorporates a salutogenetic regulatory principle of the human organism, which could be the cause of the Waldorf teachers’ reduced anxiety.

In our study, 4.6% of the teachers had contracted COVID-19, which is similar to the rate of the overall population (1). Recent studies showed that teachers do not bear an increased risk of contracting SARS-CoV-2 (16), however, the perceived risk may differ significantly. Our survey showed that nearly 35% of the teachers assess their risk of suffering severe COVID-19 as at least increased, and that a large majority fear not to receive adequate medical care due to the overloaded health system. Many patients who were hospitalized with severe COVID-19 have an underlying disease, are older or very obese (17, 18). In our study, fear was highest among the youngest and decreased with age, which strikingly contradicts the actual risk for severe COVID-19. The median age of the teachers was 43 years with under 9% reporting of having asthma or any other respiratory disease suggesting that teachers of our study do not have a higher risk of severe COVID-19 than the overall population. It can be speculated that teachers only perceive their risk as being high. In fact, our data show that teachers who assess their risk as high or moderate are significantly more anxious compared to their colleagues who perceive their risk as low, respectively. Further studies are needed to evaluate the reasons why anxiety promotes teacher’s risk perception.

Although schools were discussed to be COVID-19 infection hotspots, current evidence suggests against a major role for schools in driving the pandemic. Children and adolescents are typically less susceptible to infection (19–22) than older individuals. Additionally, previous data showed that children are rarely the index cases of clusters (8, 9, 19). In Germany, only 48 COVID-19 outbreaks were registered at schools with ≥2 cases between January and August 2020 (1), accounting for 0.5% of all reported outbreaks during this period. Further studies concluded that re-opening of schools in Germany under strict hygiene measures have not increased the number of newly confirmed SARS-CoV-2 infections (23–25). After SARS-CoV-2 infections, children usually show milder symptoms (26, 27) and up to 50% of them may remain asymptomatic (28). The actual number of infected children attending school may therefore be higher than anticipated. The concern of a large majority of our interviewed teachers that children attend school asymptomatically despite being infected is therefore justified. On the other hand, child-to-child transmission seems to be lower than intrafamily transmission and transmission from and between adults (29, 30). Also, larger school outbreaks are rather associated with higher SARS-CoV-2 positivity rates in school staff and secondary transmissions from teachers (31, 32). This is probably why many participating teachers in our study supported the notion that all colleagues should be vaccinated before returning to the classroom.

During the 2020/21 SARS-CoV-2 pandemic, many countries closed schools. Restrictions on large and prolonged social gatherings, such as educational closures, have been identified as one of the most effective non-pharmaceutical interventions (33). However, school closures may substantially disrupt the lives of children and their families and may have consequences for children’s mental health (34). In our study, most teachers believe that schools are a major contributor spreading SARS-CoV-2 and are warning to proceed with caution when re-opening schools even though previous studies convincingly showed that schools play a subordinate role spreading SARS-CoV-2 (24, 25). So, we can speculate that teachers at German schools are performing a continuous balancing act between caring about their health and well-being and their pedagogical mission. And indeed, according to latest findings of a qualitative study, teachers emphasize prioritizing and promoting health and well-being of pupils and teachers in the school learning environment (35).

Our study had limitations. Most of the participating teachers were recruited via a news magazine and teachers’ associations. Teachers who were neither members of the associations nor visit the webpage of News4teachers could therefore not be included in the study. Furthermore, teachers from vocational schools were not included. Their fear and risk perception may differ as they teach adolescents and young adults. At last, our survey was conducted from March-April 2021. During this dynamic time, Germany faced the second and third COVID-19 waves, vaccination plans changed, schools re-opened and closed depending on the state, and the government relaxed and tightened lockdown measures depending on the numbers of new cases having possibly an impact on teachers’ fear and risk perception. The proportion of female teachers in our study reflects the national gender proportion of teachers. The external validity is therefore given.

To the authors’ knowledge, this is the first study to describe teachers fear and risk perception of COVID-19 and their attitude towards vaccinations in a nationwide survey. In conclusion, this study shows a strong fear and risk perception among teachers in Germany which results in an above average vaccination intention. Further studies are needed to evaluate whether new virus variants may change infection events also in adolescents and students.

## Data Availability

The ImpfREAD surevy in German language, as well as the data that support the findings of this study are available from the corresponding author, [SW], upon reasonable request.

## Key points

- Policy makers need to balance pros and cons of the school re-opening strategy.
- Teachers’ fear of infection and risk perception was assessed using a COVID-19 specific online survey.
- A high majority (77%) of teachers in Germany fear to get infected with the SARS-CoV-2 and intended to be vaccinated against COVID-19.
- Female teachers were significantly more anxious to get infected with SARS-CoV-2. The younger the teacher, the more anxious they are and teachers who didn’t consider school re-openings to be of highest priority were 9 times more anxious.

## Acknowledgement

The authors acknowledge the contribution of Alexandra Jakubowski for her efforts in designing the survey and analyzing the data.

