## Supplemental Table S1 - S13 for "School teachers’ self-reported fear and risk perception during the COVID-19 pandemic – a nationwide survey in Germany"

**Supplementary table S1** Sources from which participants were recruited (*n=6667*).

| Source | n (%) |
|---|---|
| News4Teacher | 2803 (42.0) |
| School boards | 1101 (16.5) |
| Education and Training Association (VBE) and the Union for Education and Training (GEW) | 2173 (32.6) |
| German Teachers' Association | 330 (5.0) |
| Association of Waldorf Schools | 254 (3.8) |
| Twitter @CSozialmedizin | 4 (0.1) |
| Montessori Association | 1 (0.01) |
| Unknown | 1 (0.01) |

**Supplementary table S2** Geographical distribution of the participating teachers (*n=5128*).

| Zip code | n (%) |
|---|---|
| 00000 – 09999 | 378 (7.4) |
| 10000 – 19999 | 687 (13.4) |
| 20000 – 29999 | 658 (12.8) |
| 30000 – 39999 | 676 (13.2) |
| 40000 – 49999 | 554 (10.8) |
| 50000 – 59999 | 479 (9.3) |
| 60000 – 69999 | 363 (7.1) |
| 70000 – 79999 | 494 (9.6) |
| 80000 – 89999 | 315 (6.1) |
| 90000 – 99999 | 524 (10.2) |

**Supplementary table S3** Teachers' fear of contracting COVID-19 at school and their vaccination intention.

| Variable | n (%) |
|---|---|
| **"I'm afraid contracting coronavirus at my workplace." (*n=5936)*** | |
| I agree completely | 2526 (42.6) |
| I agree | 1807 (30.4) |
| Neither nor | 499 (8.4) |
| I don't agree | 434 (7.3) |
| I don't agree at all | 670 (11.3) |
| **"Children can carry and transmit the virus without developing COVID-19 symptoms. Is this worrying to you?" (*n=5930*)** | |
| I agree completely | 2749 (46.4) |
| I agree | 1484 (25.0) |
| Neither nor | 422 (7.1) |
| I don't agree | 464 (7.8) |
| I don't agree at all | 811 (13.7) |
| **"Do you think that schools and day-cares contribute greatly spreading of the virus?" (*n=5930*)** | |
| I agree completely | 1952 (32.9) |
| I agree | 1931 (32.6) |
| Neither nor | 781 (13.2) |
| I don't agree | 513 (8.7) |
| I don't agree at all | 753 (12.7) |

| | |
|---|---|
| **“Do you think that children contribute greatly spreading the virus in schools and day-cares?” (*n=5930*)** | |
| I agree completely | 1777 (30.0) |
| I agree | 2085 (35.2) |
| Neither nor | 843 (14.2) |
| I don’t agree | 484 (8.2) |
| I don’t agree at all | 741 (12.5) |
| **“Do you intent to receive a COVID-19 vaccination?” (*n=5850*)** | |
| Yes | 4512 (77.1) |
| No | 1019 (17.4) |
| Not sure yet | 319 (5.5) |
| **“I would like that all colleagues had received a COVID-19 vaccination before returning to classroom teaching.” (*n=5872)*** | |
| I agree completely | 2942 (50.1) |
| I agree | 1004 (17.1) |
| Neither nor | 588 (10.0) |
| I don’t agree | 332 (5.7) |
| I don’t agree at all | 1006 (17.3) |

**Supplementary table S4** Assessment of teacher’s risk perception at work.

| **Variable** | **n (%)** |
|---|---|
| **“If you agree, by whom do you feel most threatened? (Multiple answers possible)” (*n=4320*)** | |
| Students | 4224 (97.8) |
| Parents | 542 (12.6) |
| Younger colleagues (until 55 years) | 1894 (43.8) |
| Older colleagues (> 55 years) | 1308 (30.3) |
| I don’t know | 61 (1.4) |
| **“Are you a risk factor for the students or do the students put teachers at risk? (Multiple answers possible)” (*n=5840*)** | |
| Students put teachers at risk | 1113 (19.1) |
| Teachers put students at risk | 540 (9.3) |
| The risks come from both sides | 4479 (76.7) |
| A high risk only exists towards older teachers (from 60/65 years) | 685 (11.7) |
| I don’t know | 330 (5.7) |

**Supplementary table S5** Association factors for teacher's fear contracting SARS-CoV-2.

| Variable (n=6753) | Odds ratio (95% CI) | p-value |
|---|---|---|
| *Age* | 0.98 (0.97 – 0.99) | 0.0008*** |
| *Intention to get vaccinated* | | |
| Yes# | 1 | |
| No | 0.02 (0.02 – 0.03) | <0.0001*** |
| Not sure | 0.17 (0.12 – 0.23) | <0.0001*** |
| *Gender* | | |
| Male# | 1 | |
| Female | 1.92 (1.50 – 2.45) | <0.0001*** |
| Diverse | 0.63 (0.12 – 3.25) | 0.5818 |
| *Schools* | | |
| Elementary schools# | 1 | |
| Secondary schools (Hauptschulen) | 1.20 (0.66 – 2.20) | 0.5442 |
| Secondary schools (Realschulen) | 1.12 (0.74 – 1.68) | 0.5947 |
| High schools | 0.96 (0.68 – 1.35) | 0.8014 |
| Comprehensive schools | 0.99 (0.68 – 1.45) | 0.9760 |
| Schools for children with special needs | 0.63 (0.42 – 0.95) | 0.0264* |
| Waldorf schools | 0.38 (0.24 – 0.62) | <0.0001*** |
| Montessori schools | 0.57 (0.11 – 2.92) | 0.5006 |
| Boarding schools | 3.17 (0.01 – 1204.81) | 0.7033 |
| Privat schools | 0.79 (0.39 – 1.60) | 0.5136 |
| Language schools | 3.15 (0.56 – 1.78) | 0.1945 |
| *Opening schools as highest priority* | | |
| I agree completely# | 1 | |
| I agree | 1.63 (1.26 – 2.09) | 0.0002*** |
| Neither nor | 2.04 (1.53 – 2.72) | <0.0001*** |
| I don't agree | 4.87 (3.49 – 6.80) | <0.0001*** |
| I don't agree at all | 8.94 (4.69 – 17.05) | <0.0001*** |
| *Risk perception of getting COVID-19* | | |
| High# | 1 | |
| Moderate | 0.12 (0.08 – 0.17) | <0.0001*** |
| Little | 0.03 (0.02 – 0.04) | <0.0001*** |
| No | 0.01 (0.00 – 0.02) | <0.0001*** |
| *Subjects taught* | | |
| German language, no# | 1 | |
| German language, yes | 0.96 (75 – 1.23) | 0.7542 |
| Mathematics, no# | 1 | |
| Mathematics, yes | 0.92 (0.72 – 1.16) | 0.4611 |
| Science, no# | 1 | |
| Science, yes | 0.96 (0.76 – 1.22) | 0.7263 |
| „Science at elementary schools", no# | 1 | |
| „Science at elementary schools ", yes | 0.79 (0.60 – 1.03) | 0.0840 |
| Foreign languages, no# | 1 | |
| Foreign languages | 0.80 (0.64 – 0.99) | 0.0424* |
| Sports, no# | 1 | |
| Sports, yes | 0.90 (0.69 – 1.18) | 0.4593 |
| Music, no# | 1 | |
| Music, yes | 1.34 (1.00 – 1.81) | 0.0501 |
| Others, no# | 1 | |
| Others, yes | 1.08 (0.88 – 1.33) | 0.4447 |

Multivariable analysis for association factors of teacher's fear getting infected with SARS-CoV-2.

#references; p values (two-sided, paired t test) for association between references and respective answers. *p value <0.05; *** p value <0.001

**Supplementary table S6** Teachers' perception towards the student's parents and COVID-19 precautions.

| Variable | n (%) |
|---|---|
| **"Many parents are too careless with corona." (*n=5926*)** | |
| I agree completely | 978 (16.5) |
| I agree | 1812 (30.6) |
| Neither nor | 1485 (25.1) |
| I don't agree | 1052 (17.8) |
| I don't agree at all | 599 (10.1) |
| **"Many parents only think about themselves and childcare." (*n=5925*)** | |
| I agree completely | 1003 (16.9) |
| I agree | 1674 (28.3) |
| Neither nor | 1500 (25.3) |
| I don't agree | 1180 (19.9) |
| I don't agree at all | 568 (9.6) |
| **"I give in to the parents' insistence on additional hygiene measures (group divisions, setting up disinfection dispensers, etc.) ...." (*n=5820*)** | |
| Always | 553 (9.5) |
| Sometimes | 1140 (19.6) |
| Rarely | 950 (16.3) |
| Never | 727 (12.5) |
| I don't know | 2450 (42.1) |
| **"The requirements regarding the implementation of the COVID-19 precautions of the state put me under pressure." (*n=5931*)** | |
| I agree completely | 1443 (24.3) |
| I agree | 2020 (34.1) |
| Neither nor | 923 (15.6) |
| I don't agree | 1225 (20.7) |
| I don't agree at all | 320 (5.4) |
| **"I feel I cannot fullfil the expectations of parents and the state with regard to the implementation of the COVID-19 precautions while considering children's needs." (*n=5907*)** | |
| I agree completely | 1077 (18.2) |
| I agree | 1826 (30.9) |
| Neither nor | 1502 (25.4) |
| I don't agree | 1069 (18.1) |
| I don't agree at all | 433 (7.3) |

**Supplementary table S7** Frequencies teachers are getting tested.

| Variable | n (%) |
|---|---|
| **"How often do you get tested for a corona infection?" (*n=5794*)** | |
| Once a week | 1917 (33.1) |
| Once a month | 164 (2.8) |
| According to the risk situation | 2441 (42.1) |
| Never before | 1272 (22.0) |

**Supplementary table S8** Teachers' opinion on COVID-19 becoming seasonal.

| Variable | n (%) |
|---|---|
| **"I can imagine that COVID-19 could also become a seasonal disease like influenza due to mutations in the corona virus." (*n=5712*)** | |
| I agree completely | 1483 (26.0) |
| I agree | 2943 (51.5) |
| Neither nor | 983 (17.2) |
| I don't agree | 213 (3.7) |
| I don't agree at all | 90 (1.6) |

**Supplementary table S9** Teachers' preferences for vaccines. (*n=4505*)

| Variable | n (%) |
|---|---|
| **"If you had the choice, which vaccine would you prefer? (Multiple answers possible)"** | |
| BioNTech | 3249 (72.1) |
| Moderna | 1955 (43.4) |
| AstraZeneca** | 989 (22.0) |
| Janssen | 110 (2.4) |
| Sputnik V | 235 (5.2) |
| Johnson&Johnson | 693 (15.4) |
| I cannot judge that | 355 (7.9) |
| I don't mind any | 1025 (22.8) |

**It should be noted, that much of the survey was conducted before the sinus vein thromboses became known as an adverse event of vaccination with the AstraZeneca vaccine. As of April 1st, 2021, the Standing Committee on Vaccination (STIKO) restricted the AstraZeneca vaccine to the age group ≥ 60 years.

**Supplementary table S10** Teachers' reasons for or against COVID-19 vaccination.

| Variable | n (%) |
|---|---|
| **"What are the reasons you intent to get vaccinated? (Multiple answers possible)" (*n=4505*)** | |
| For my own safety to be protected against COVID-19 | 4367 (96.9) |
| To be able to return to work quickly | 1190 (26.4) |
| Protection for family and friends | 4123 (91.5) |
| Protection for students | 3367 (74.7) |
| Protection for others | 3499 (77.7) |
| Vaccination certificate for more freedom to travel | 1141 (25.3) |
| To spend leisure time as per my wish | 1504 (33.4) |
| Special rights for vaccinated people | 493 (11.0) |
| Reduction of lockdown measures | 2407 (53.4) |
| No reason given | 7 (0.2) |

| “What are the reasons you intent NOT to get vaccinated? (Multiple answers possible)” (*n=1008*) | |
|---|---|
| I already had COVID-19 disease and recovered | 86 (8.5) |
| Concerns about vaccine reaction and adverse events | 769 (76.3) |
| Concerns about as yet unknown long-term effects | 881 (87.4) |
| I do not trust new vaccines | 525 (52.1) |
| Vaccines not yet properly tested | 781 (77.5) |
| I do not trust the manufacturers | 494 (49.0) |
| I do not believe in COVID-19 | 71 (7.0) |
| I have a pre-existing condition (contraindication/increased risk from vaccination) | 95 (9.4) |
| I am pregnant / I am breastfeeding | 31 (3.1) |
| No reason given | 42 (4.2) |

**Supplementary table S11** Teachers’ opinion on compulsory vaccinations.

| Variable | n (%) |
|---|---|
| **“I support compulsory vaccination against COVID-19, which is being discussed by politicians.” (*n=5782*)** | |
| I agree completely | 1137 (19.7) |
| I agree | 1372 (23.7) |
| Neither nor | 973 (16.8) |
| I don’t agree | 960 (16.6) |
| I don’t agree at all | 1340 (23.2) |
| **“In general, I support compulsory vaccinations as against measles also for other infectious diseases.” (*n=5785*)** | |
| I agree completely | 1333 (23.0) |
| I agree | 1486 (25.7) |
| Neither nor | 947 (16.4) |
| I don’t agree | 834 (14.4) |
| I don’t agree at all | 1185 (20.5) |

**Supplementary table S12** Teachers’ attitude towards vaccinations.

| Variable | n (%) |
|---|---|
| **“Do you have regular checks to ensure that you have received all recommended vaccinations?” (*n=5772*)** | |
| Ja | 4016 (69.6) |
| Nein | 1756 (30.4) |
| **“Do you consider the recommendations of the Standing Committee on Vaccination (STIKO) in Germany to be ...” (*n=5772*)** | |
| Appropriate | 3551 (61.5) |
| Too little | 414 (7.2) |
| Exaggerated | 937 (16.2) |
| I don’t know | 870 (15.1) |

**Supplementary table S13** Teachers' opinion on COVID-19 vaccine allocation and school re-openings.

| Variable | n (%) |
|---|---|
| **"I think a prioritized allocation of the COVID-19 vaccine is appropriate." (*n=5797*)** | |
| I agree completely | 1622 (28.0) |
| I agree | 2489 (43.0) |
| Neither nor | 959 (16.5) |
| I don't agree | 374 (6.4) |
| I don't agree at all | 353 (6.1) |
| **"I think that the new classification of teachers into group - 2 (high priority) is absolutely justified." (*n=5799*)** | |
| I agree completely | 2592 (44.7) |
| I agree | 1640 (28.3) |
| Neither nor | 812 (14.0) |
| I don't agree | 351 (6.1) |
| I don't agree at all | 404 (7.0) |
| **„I think that re-opening up schools is an absolute priority?" (*n=5824*)** | |
| I agree completely | 1525 (26.2) |
| I agree | 1544 (26.5) |
| Neither nor | 1104 (19.0) |
| I don't agree | 1271 (21.8) |
| I don't agree at all | 380 (6.5) |
